# Global Reporting Patterns of Suspected Adverse Drug Reactions and Their Relationship with a Country’s Socioeconomic Status- Insights from VigiBase Data- Study Protocol

**DOI:** 10.1101/2025.02.14.25322050

**Authors:** Nisha Dhakal, Daniele Sartori, Anna-Riia Holmström

## Abstract

**Introduction:** Pharmacovigilance is vital for detecting Adverse Drug Reactions (ADRs), with Individual Case Safety Reports (ICSRs) serving as the primary source, despite growing interest in other data sources like electronic health records and social media. However, ICSR reporting rates remain low in low- and middle-income countries due to weak pharmacovigilance systems. Furthermore, the impact of socioeconomic factors such as health, education, and income on reporting rates has not been studied on a global scale previously.

**Aims:** To compare global ICSR reporting patterns and quantify potential differences in their characteristics across pre-defined socioeconomic strata.

**Methods:** In this observational descriptive quantitative study, numbers of ICSRs will be obtained from VigiBase for members of the World Health Organization Programme for International Drug Monitoring, from 01/01/2010 to 31/12/2022. Yearly total population data and Inequality-adjusted Human Development Index (IHDI) values will be obtained from the United Nations Development Programme (UNDP) for all countries, for the same interval. Yearly ICSR reporting rates for each country will be calculated by dividing the total ICSRs submitted to VigiBase each year, and median reporting rates computed across the study period. Similarly, the median IHDI and its dimensional indices (Health, Education, and Income) will be calculated for each country. Countries will be categorized into four human development groups based on the UNDP classification: low, medium, high, and very high. To assess the relationship between IHDI (and its dimensions) and reporting rates, a linear regression model will be applied, with adjustments made as needed based on the data characteristics. VigiPoint will also be used to compare the relative frequency of covariates between reports of low, medium, and high IHDI, against those submitted to VigiBase by very high IHDI countries. VigiPoint identifies key features for review using shrunk log odds ratios, highlighting the overrepresented characteristics of ICSRs. We will present descriptive and statistical analyses in tables and scatter plots will be used to examine the relationship between ICSR reporting rates and IHDI (and its dimensions), using the calculated median values for IHDI (and its dimensions). Trendlines will highlight temporal changes in population, IHDI, and dimensional indices.

**Discussion:** The findings will offer insights into how socioeconomic factors affect reporting practices and may guide targeted interventions to enhance global drug safety monitoring.

## Introduction

Pre-clinical and clinical studies represent critical phases in the development of an investigational drug, which are essential for evaluating its safety profile prior to market release. However, such studies include a limited number of study subjects for limited time and cannot determine the in-depth safety profile of a new drug (Inácio, Cavaco, and Airaksinen 2017). Pharmacovigilance is “the science and activities relating to the detection, assessment, understanding, and prevention of adverse effects or any other possible drug-related problems” (World Health Organization 2002). This field plays a crucial role in detecting rare Adverse Drug Reactions (ADRs) throughout the life cycle of a medicinal product. According to WHO “An adverse drug reaction (ADR) is ‘a response to a medicine which is noxious and unintended, and which occurs at doses normally used in man”(World Health Organization 2002).ADRs significantly affect diverse global populations, contributing to morbidity and mortality, particularly among vulnerable groups such as the elderly, children, and individuals in low-income communities. (Le Louët and Pitts 2023). Therefore, pharmacovigilance gathers information regarding ADRs from Individual Case Safety Reports (ICSRs), filling the evidence gap from pre-marketing safety data (Aronson 2023). ICSR refers to the format and content for reporting one or several suspected ADRs concerning a medicinal product that occurs in a single patient at a specific time (European Medicine Agency 2014). The systematic collection and analysis of ADRs began after the thalidomide tragedy in 1961 (van Grootheest 2003). The WHO (World Health Organization) Programme for International Drug Monitoring (PIDM) began in 1968, and a decade later, the Uppsala Monitoring Centre (UMC) was founded to handle its operational functions. UMC was designated as a WHO Collaborating Centre for International Drug Monitoring to serve as a global hub for detecting and monitoring ADRs. To date, UMC manages VigiBase, the WHO global database of adverse event reports for medicines and vaccines, collecting ICSRs from members of the WHO PIDM (MARIE LINDQUIST and I. RALPH EDWARDS 2001). The WHO PIDM has two types of member countries: full members and associate members. Full members of the WHO PIDM are national centers that have completed the application process and met all reporting requirements, allowing them to fully engage in the pharmacovigilance network. Associate members are those whose applications have been accepted but are still in a transitional phase, receiving support from WHO and UMC while working towards full membership by submitting ICSRs and ensuring compliance with reporting standards (Uppsala Monitoring Centre, n.d.). However, reporting rates of ADRs remain notably low in low- and middle-income countries due to weak regulatory enforcement and limited awareness of pharmacovigilance (Olsson et al. 2010).

To our knowledge, only one global study has studied ADRs reporting rates in VigiBase data, with a focus on exploring the connection between these reporting rates and national income levels, categorized by the World Bank as low-income, lower-middle-income, upper-middle-income, and high-income (Aagaard et al., 2012). Another socioeconomic index, the Human Development Index (HDI), was developed to emphasize that a country’s development should be assessed based on the well-being and capabilities of its people. It incorporates additional parameters, namely health and education, rather than relying solely on economic growth (UNDP n.d.). Few studies have shown the correlation between HDI, and the economic growth of countries (Elistia & Syahzuni, 2018). However, few countries do not show such a correlation between economic growth and HDI (Pekarčíková & Prachařová, 2023). Consequently, it can be inferred that countries with high Gross National Income (GNI) per capita may still exhibit low human development outcomes. For example, in the United Nations Development Programme’s (UNDP) Human Development rankings, Switzerland ranks 1st with a GNI per capita of $69,433, while Qatar, despite a higher GNI per capita of $95,944, ranks 40th in HDI (UNDP 2024).

This suggests that the HDI can be helpful in examining how various socioeconomic factors influence a country’s ADR reporting rates to VigiBase. One significant factor is the level of education among patients, which is frequently cited as a reason for the underreporting of ADRs (Costa et al. 2023). Additionally, health expenditure and the income level of a country also affect these reporting rates (Vaidya et al., 2010). Therefore, it can be argued that income status, education level, and residents’ access to healthcare services represent essential global targets for advancement in pharmacovigilance (Costa et al. 2023; Vaidya et al. 2010).

In this study, we shall use the Inequality-adjusted Human Development Index (IHDI) as a socioeconomic index, which adjusts the HDI to account for inequalities in life expectancy, education, and income within a country (United Nations Development Programme 2023). This approach aims to reflect a more accurate measure of human development by factoring in inequality within a country. We seek to expand this by examining how overall human development, adjusted for inequality, may impact that country’s pharmacovigilance practices with a focus on reporting the rate of suspected ADRs as well as the differences in the patterns of ICSRs in different IHDI levels of countries.

## Aims and Objectives

The aim of study is to compare ICSR reporting patterns and quantify potential differences in their characteristics across pre-defined socioeconomic strata of countries at the global level.

The specific objectives are to study country-level correlations between IHDI (and its dimensional indices) and reporting rates, and to investigate differences in reports’ characteristics across IHDI groupings.

## Materials and Methods

### Data Sources

#### VigiBase

This observational descriptive quantitative study will use ICSRs submitted to VigiBase by full member countries of the WHO PIDM. Adverse events in VigiBase are coded to the latest version of the Medical Dictionary for Regulatory Activities (MedDRA), while medicinal products to WHODrug Global (Lagerlund et al. 2020). Numbers of deduplicated ICSRs will be retrieved for each country from VigiBase, irrespective of the involvement of a medicinal product (suspected, interacting, or concomitant).

#### UNDP Human Development Reports Data Center

IHDI values, along with the corresponding values for its three dimensions and population data for the study period, will be sourced from the UNDP Human Development Report Data Center, where IHDI data is available, but the dimensional indices are not, we shall calculate these indices using data and the tool provided on the UNDP site, following the methodologies outlined in the technical notes provided by UNDP (United Nations Development Programme 2023)

The HDI provides a summary of a country’s achievements in three dimensions of human development: living a long and healthy life, access to education, and having a decent standard of living. To calculate the HDI, two main steps are followed. First, dimension indices for health, education, and income are created using predefined minimum and maximum values (goalposts) to standardize the indicators to a scale from 0 to 1. For health, life expectancy at birth is used; for education, both expected and mean years of schooling are considered; and for income, gross national income (GNI) per capita is transformed using its natural logarithm. The education index is the arithmetic mean of the two schooling indicators. In the second step, the overall HDI is calculated as the geometric mean of the three-dimension indices (United Nations Development Programme 2023).

The formula for calculating the HDI is :

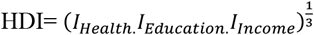

The IHDI modifies the HDI to account for inequality in the distribution of its dimensions across the population. It begins by estimating inequality in these dimensions using the Atkinson inequality measure, which compares the geometric mean (g) and arithmetic mean (μ) of each dimension’s distribution. The resulting inequality measure (Ax) is calculated for each dimension, noting that for mean years of schooling, a value of one year is added to avoid zero values. Next, the dimension indices are adjusted for inequality by multiplying the original HDI indices (Ix) by (1 - Ax). Followingly, the IHDI is computed as the geometric mean of these adjusted indices. This allows the IHDI to reflect the impact of inequality on overall development (United Nations Development Programme 2023)

Formula for IHDI Calculation:

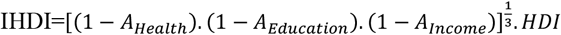

Where, A= Calculated inequality in respective distributions

#### Inclusion and Exclusion Criteria

This study will include ICSRs submitted to VigiBase by full members of the WHO PIDM between the 1^st^ of January 2010, and the 31^st^ December 2022. ICSR submissions outside this timeframe will be excluded, as the IHDI was introduced in 2010, with the most recent data available up to 2022. Additionally, ICSRs related to COVID-19 vaccines will be excluded to minimize the impact of the large influx of ICSRs on reporting patterns resulting from the pandemic. ICSRs from countries that are no longer members of the WHO PIDM will also be excluded, along with those countries lacking IHDI values for the entire study period, as IHDI is a key variable in this study.

#### Categorization of countries

The countries in this study will be classified into four human development groups based on their median IHDI values over the study period, following the United Nations Development Programme (UNDP) classification. The groups are defined as follows: very high (IHDI above 0.800), high (IHDI between 0.700 and 0.799), medium (IHDI between 0.550 and 0.699), and low (IHDI below 0.550). The IHDI values range from 0 to 1 (United Nations Development Programme, 2023). A similar classification will be applied to countries based on their dimensional index values.

### Data Analysis

#### Descriptive Statistics

Annual ICSR reporting rates for each country will be calculated by dividing the total number of ICSRs submitted per year by that year’s population. From these annual rates, a median reporting rate will be calculated for each country across the entire study period. Likewise, the median IHDI and dimensional indices for each country will be derived over the study period. Countries will be categorized into four human development groups according to median IHDI values and dimensional indices, based on the UNDP human development classification.

A table will present each country’s ICSR reporting rates, population data, IHDI values, and dimensional indices over the study period. To examine the relationship between ICSR reporting rates and IHDI (along with its dimensional indices), scatter plots will be generated. In these scatter plots, the X-axis will represent the median reporting rate, and the Y-axis will represent the median IHDI (and dimensional indices). Additionally, trendlines will depict temporal changes in population, IHDI values, and dimensional indices throughout the study period.

### Statistical Analysis

To assess the relationship between the independent variables (IHDI and its dimensions) and the dependent variable (ADR reporting rate), a linear regression model is initially planned; however, the model may be adjusted later based on the characteristics of the data.

All statistical analysis and visualization will be performed using Python 3.12.4.

### Characteristics of ICSRs across different IHDI groups (high, medium, and low) compared to a very high IHDI group

VigiPoint is a data exploration tool in pharmacovigilance that identifies associations between subjects of interest and key covariates in ADR reports available in VigiBase. Mathematically, an odds ratio with adaptive statistical shrinkage compares a data subset of interest to one or more comparator data sets, identifying statistical differences in the relative frequencies of their respective covariates. In this study, subsets of interest are low IHDI group, medium IHDI group, and high IHDI group and the comparator data set is very high IHDI group, and the covariates of interest for analysis will include Age group, Completeness(A metric that quantifies the amount of clinically relevant information contained in ICSRs (Uppsala Monitoring Centre n.d.-b)), Reported Fatality, Notifier of ICSRs, Patient sex, Report Type, Seriousness(An ICSR is classified as serious if it meets one or more of the following criteria: results in death, is life-threatening, requires or prolongs hospitalization, leads to persistent or significant disability/incapacity, causes a congenital anomaly/birth defect, or is deemed medically important by a healthcare professional (ICH Expert Working Group 2001)), Group of reported drugs according to ATC third level (SpATClvl3), Substance name of drugs reported as suspected (CoSuspDrug), Events coded to high-level terms in MedDRA (MedDRAHLT), and to preferred terms (MedDRAPT).

Results from the statistical analysis will be presented in a table.

## Discussion/Conclusion

To our knowledge, this study will be the first to consider both GNI per capita and IHDI and may reveal links between reporting patterns and overall human development, beyond a country’s economic status. It may also reveal differences in characteristics of ICSRs across strata of IHDI, identifying classes of medicinal products or groups of adverse effects that are more likely to be reported in sets of countries. Overall, this study can contribute to furthering the understanding of global pharmacovigilance practices and pitfalls, that may be of help to policymakers to target interventions to optimize surveillance of medicinal products where necessary. A report from this study will be presented as MSc thesis work at the University of Helsinki, Finland, and will be submitted for peer-reviewed publication in a suitable scientific journal.

## Limitations

One limitation of this study is that the number of ICSRs submitted to VigiBase may differ from the counts reported by national pharmacovigilance centers in individual countries. This discrepancy could influence the visualization of the relationship between reporting rates and independent factors. However, the large volume of reports in VigiBase may help mitigate this potential statistical issue. Additionally, we also excluded all ICSRs related to COVID-19 vaccines to reduce bias, given that the study period overlaps with the SARS-CoV-2 pandemic. The use of other supplementary medications during this time may still influence the relationship between reporting rates and socioeconomic factors. Furthermore, the availability and use of different classes of medications can vary across IHDI groups, which may affect this association and contribute to differences in the characteristics of reports submitted across these groups.

## Research Ethics and Data Management

Ethical board review is not required for this study, since it will use de-identified VigiBase data. However, an agreement for data handling between the researcher and UMC will be established, as UMC treats the data as sensitive and strictly manages access to it.

## Disclosures

Nisha Dhakal (Student, master’s degree in Pharmaceutical Research, Development and Safety, Faculty of Pharmacy, University of Helsinki) has no relevant conflicts of interest.

Anna-Riia Holmström (Assistant professor at University of Helsinki) has no relevant conflicts of interest.

Daniele Sartori (Pharmacovigilance Scientist at Uppsala Monitoring Centre) has no relevant conflicts of interest.

The information that will be presented in this study will not represent the opinion of the UMC or the World Health Organization. According to the VigiBase Caveat Document, “the information [in the database] comes from a variety of sources, and the probability that the suspected adverse effect is drug-related is not the same in all cases”.

## Data Availability

All data produced in the present study are available upon reasonable request to the authors

